# Health-related quality of life in individuals with syndromic autism spectrum disorders

**DOI:** 10.1101/2020.06.10.20127951

**Authors:** Corneliu Bolbocean, Fabiola N. Andújar, Maria McCormack, Bernhard Suter, J. Lloyd Holder

## Abstract

**BACKGROUND:** Children with autism have a significantly lower quality of life compared with their neurotypical peers. While multiple studies have quantified the impact of autism on health-related quality of life (HRQoL) through standardized surveys such as the PedsQL, none have specifically investigated the impact of <u>syndromic</u> autism spectrum disorder on children’s HRQoL or on family quality of life. Here we evaluate HRQoL in children diagnosed with three syndromic Autism Spectrum Disorders (ASDs): Phelan-McDermid syndrome (PMD), Rett syndrome (RTT), and *SYNGAP1*-related intellectual disability (*SYNGAP1*-ID).

**METHODS:** A standardized online Pediatric Quality of Life Inventory (PedsQL 4.0) survey and the Beach Center Family Quality of Life Scale (FQOL) were administered to caregivers of children with PMD (n= 213), RTT (n= 148), and *SYNGAP1*-ID (n= 30). The PedsQL 4.0 measures health-related quality of life in four dimensions: physical, emotional, social and school. The Beach Center Family Quality of Life Scale measures five dimensions: family interaction, parenting, emotional well-being, physical/material well-being and disability-related support.

**RESULTS:** For the PedsQL, the most severely impacted dimension in children with syndromic autism was physical functioning. In comparing individual dimensions among the genetically-defined syndromic autisms, individuals with RTT had significantly worse physical functioning, emotional and school scores than PMD. This finding is congruent with the physical regression typically associated with Rett syndrome. Strikingly, syndromic autism results in worse quality of life than other chronic disorders including idiopathic autism.

**CONCLUSIONS:** The reduced HRQoL for children with syndromic autism spectrum disorders relative to other chronic childhood illnesses, likely reflects their lack of targeted therapies. This study demonstrates the utility of caregiver surveys in prioritizing phenotypes, which may be targeted as clinical endpoints for genetically defined ASDs.

**CONTRIBUTORS’ STATEMENT:** Dr. Bolbocean conceptualized and designed the study, designed the data collection instrument, collected data, performed data analysis, wrote and edited the manuscript.

Ms. Andujar performed initial data analysis, drafted the initial manuscript and edited the manuscript.

Ms. McCormack performed data analysis and edited the manuscript.

Dr. Suter conceptualized and designed the study and made critical edits to the manuscript.

Dr. Holder conceptualized and designed the study, designed the data collection instrument, performed data analysis, wrote and edited the manuscript.

**Table of contents summary:** In this study, we determine the impact of genetically-defined syndromic autism spectrum disorders on their health-related quality of life.

**What’s known on this subject:** Children with neurodevelopmental disorders, including autism, have severely impaired health-related quality of life. Systematic measurement of HRQoL in children with neurodevelopmental disorders through standardized instruments provides a holistic understanding of disease impact and therapeutic endpoint for clinical trials.

**What this study adds:** This study defines the impact of three genetically defined autism spectrum disorders: Rett syndrome, Phelan-McDermid syndrome and *SYNGAP1*-related Intellectual Disability, on health-related quality of life. We find significantly greater impairment for syndromic ASDs than other neurodevelopmental disorders.

## INTRODUCTION

The diagnosis of autism spectrum disorders (ASDs) has increased dramatically over the past decade[1]. The current U.S. prevalence is 1 in 59 children (1.7%) by the age of 8, with boys diagnosed 4 to 5 times more often than girls[2]. Diagnostic criteria for ASDs have been revised in the most recent Diagnostic and Statistical Manual of Mental Disorders, 5^th^ Edition (DSM-5)[3]. Autism is diagnosed if the two main criteria are met: persistent deficits in social communication and restrictive or repetitive patterns of behavior. Importantly, two qualifiers must also be present: the onset of symptoms must be early in development and the symptoms, particularly delayed communication, must be greater than expected for impairments in other developmental domains.

Autism spectrum disorders are defined as either syndromic or non-syndromic. Children diagnosed with non-syndromic ASD meet the DSM-5 criteria without other significant somatic or neurologic manifestations. This constitutes the majority of individuals diagnosed with autism. In contrast, syndromic ASDs are diagnosed in individuals meeting DSM-5 criteria but also manifesting somatic symptoms or additional neurologic phenotypes. The somatic symptoms may be facial or other physical dysmorphic features or more complex congenital organ abnormalities. Neurologic diagnoses commonly co-occurring with ASD include intellectual disability and epilepsy[4, 5]. Children with a clinical diagnosis of syndromic autism are more likely to have a genomic abnormality than individuals with non-syndromic ASD[6, 7].

Prolonged and multidimensional clinical issues associated with syndromic ASDs are linked to negative lifelong health and socio-economic impact of the affected child and caregivers[8, 9]. Thus, understanding the Health-Related Quality of Life (HRQoL) in children with syndromic ASD and their caregivers is critical to design clinically effective and economically viable interventions for these children.

Health-related quality of life is a multidimensional concept designed to directly measure an individual’s states related to physical, psychological, and social cognitive aspects of life[10]. Literature on HRQoL in children has identified a lower HRQoL in autism relative to other chronic disorders. However, very few studies have examined the HRQoL in pediatric patients diagnosed with syndromic autism. One such study reported that children with Fragile X Syndrome have significantly impaired health-related quality of life with cognitive function most affected[11]. However, HRQoL for other genetic disorders causing syndromic autism have not been widely reported.

This study aims to evaluate the HRQoL of individuals with three syndromic autism disorders: Rett syndrome (RTT), Phelan-McDermid syndrome (PMD) and *SYNGAP1*-related Intellectual Disability (*SYNGAP1*-ID), using the PedsQL 4.0 Inventory (see Appendix for more details of clinical phenotypes associated with each disorder). By combining data from multiple genetic disorders that cause syndromic autism for the first time, we obtain an overview of the impact of syndromic autism on HRQoL. Moreover, we are able to compare HRQoL between each genetically-defined autism and with other chronic childhood illnesses to discover the severe impact of these disorders on the affected children and their families.

## METHODS

### Participants

This study employed a cross-sectional design. Participants included 391 families with children clinically diagnosed with syndromic ASD; 213 (54.5%) of the participants were diagnosed with PMD, 148 (37.9%) with RTT, and 30 (7.7%) with *SYNGAP1*-ID. Inclusion criteria consisted of children 2-18 years of age with an ASD diagnosis of PMD, RTT or *SYNGAP1*-ID. For recruitment, an email providing access to an online Qualtrics survey was provided to families with self-identified interest in participation through the following organizations: Phelan-McDermid Syndrome Foundation, RettSyndrome.org, or Bridge the Gap: *SYNGAP1* Education and Research Foundation. Patients were recruited from all over the country.

### Procedure

Approval for the study was obtained from Baylor College of Medicine’s Institutional Review Board. All data was ascertained through caregiver self-report questionnaires. For the purposes of our study, the Pediatric Quality of Life Inventory™ version 4.0 (PedsQL 4.0) was administered as a quantifiable tool with which to assess the health-related quality of life in children diagnosed with the syndromic ASDs of interest. The Beach Center Family Quality of Life Scale (FQOL) questionnaire was also administered and assessed parents’ life satisfaction with their nuclear family. The surveys were estimated to take approximately 15 minutes to complete. They were anonymous, and the only other information caregivers were requested to provide was their child’s diagnosis. No compensation was provided.

### PedsQL 4.0

The PedsQL 4.0 is one of the most widely studied assessments of child QOL reports[10-14]. As an assessment instrument, it provides a modular approach in measuring health-related quality of life (HRQoL) in healthy children as well as those with acute or chronic conditions across four domains: physical, emotional, social, and school functioning. These domains have been identified as core dimensions of health by the World Health Organization[13]. These dimensions are reflected in the survey as four scales comprising physical (8 items), emotional (5 items), social (5 items), and school functioning (5 items). From these four core scales, a total quality of life score (based on all items) was calculated[14]. The survey consisted of 23 items in the format: “In the past ONE month, how much of a problem has your child had with…,” rated on a five-point Likert scale: (*0 = Never a problem, 1 = Almost never a problem, 2 = Sometimes a problem, 3 = Often a problem, 4 = Almost always a problem)*.

All items in the PedsQL were reverse scored and linearly transformed to a 0 to 100 scale, with higher scores indicative of greater HRQoL per standard reporting practice. Consistent with standard scoring practices, scores were transformed to (0 = 100, 1 = 75, 2 = 50, 3 = 25, and 4 = 0), and means were computed for each domain. To calculate the total scale score for each syndromic autism, we computed the mean as the sum of all items across all modules. We focused our subsequent analysis on the PMD and RTT data sets due to the relatively small number of *SYNGAP1*-ID participants.

### FQOL

The Beach Center Family Quality of Life (FQOL) Scale is a standardized instrument that is a validated measure of quality of life for families with children[15-17]. The Beach Center FQOL Scale was administered as a 25-item inventory utilizing satisfaction as the primary response format with 5 dimensions: family interaction, parenting, emotional well-being, physical/material well-being, and disability-related support. Response options were rated on a five-point Likert scale: (*1 = very dissatisfied, 2 = dissatisfied, 3 = neither satisfied nor dissatisfied, 4 = satisfied, and 5 = very satisfied*).

### Analysis

Participants were divided into three groups based on medical diagnosis. We implemented parametric and non-parametric methods to test for mean differences across PedsQL and FQOL dimensions between groups (Table 1 and 2). We removed *SYNGAP1*-ID participants from further analysis due to relative small sample size. We further computed Cronbach’s alpha and conducted principal components analyses for the PedsQL and FQOL, separately for Rett and Phelan-McDermid syndrome patients and their respective caregivers. All analyses were conducted in STATA 15.

**Table 1.** PedsQL Summary.

| **DOMAINS** | **Combined**<br>**Syndromic**<br>**ASDs**<br>**N=391**<br>**(St. Dev.)** | **PMD**<br>**N=213**<br>**(St. Dev.)** | **RTT**<br>**N=148**<br>**(St. Dev.)** | **SYNGAP1-ID**<br>**N=30**<br>**(St. Dev.)** | **Kruskal-**<br>**Wallis H**<br>**Test**<br>**p-value** |
| --- | --- | --- | --- | --- | --- |
| **PHYSICAL** | 33.55<br>(11.09) | 42.09<br>(15.51) | 19.64<br>(9.35) | 38.92<br>(9.94) | 0.001 |
| 1. Problems with walking more than one block | 45.44<br>(19.45) | 61.03<br>(15.53) | 23.64<br>(4.43) | 51.67<br>(3.16) | 0.001 |
| 2. Problems with running | 33.44<br>(16.59) | 47.64<br>(8.40) | 15.20<br>(3.39) | 37.5<br>(4.85) | 0.001 |
| 3. Problems with participating in sports activities or exercise | 27.21<br>(10.20) | 34.43<br>(3.55) | 15.54<br>(2.93) | 31.67<br>(5.05) | 0.001 |
| 4. Problems with lifting something heavy | 29.76<br>(14.38) | 37.32<br>(4.15) | 13.17<br>(4.16) | 38.79<br>(2.28) | 0.001 |
| 5. Problems with taking a bath or shower by him or herself | 19.06<br>(6.42) | 19.95<br>(4.86) | 12.24<br>(4.80) | 25.00<br>(8.16) | 0.046 |
| 6. Problems with doing chores around the house | 21.53<br>(9.97) | 24.06<br>(1.79) | 10.54<br>(4.19) | 30.00<br>(5.09) | 0.001 |
| 7. Problems with having hurts or aches | 46.88<br>(11.35) | 54.97<br>(7.05) | 33.90<br>(6.05) | 51.79<br>(4.62) | 0.001 |
| 8. Problems with low energy level | 45.09<br>(12.20) | 57.34<br>(7.05) | 32.93<br>(6.39) | 45.00<br>(4.24) | 0.001 |
| **EMOTIONAL** | 55.13<br>(10.24) | 62.14<br>(10.96) | 51.59<br>(7.67) | 51.65<br>(14.14) | 0.001 |
| 1. Problems with feeling afraid or scared | 56.63<br>(9.23) | 67.18<br>(11.19) | 52.73<br>(8.54) | 50.00<br>(4.36) | 0.001 |
| 2. Problems with feeling sad or blue | 56.66<br>(6.84) | 64.33<br>(9.75) | 51.19<br>(9.80) | 54.46<br>(6.66) | 0.001 |
| 3. Problems with feeling angry | 51.83<br>(9.74) | 59.38<br>(8.29) | 55.27<br>(10.38) | 40.83<br>(7.62) | 0.001 |
| 4. Problems with trouble sleeping | 41.03<br>(3.60) | 45.18<br>(5.14) | 39.11<br>(5.52) | 38.79<br>(2.59) | 0.169 |
| 5. Problems with worrying about what will happen to him or her | 69.48<br>(8.52) | 74.63<br>(22.09) | 59.64<br>(9.68) | 74.17<br>(5.39) | 0.001 |
| **SOCIAL** | 39.83<br>(24.47) | 38.45<br>(21.51) | 37.71<br>(29.57) | 43.34<br>(23.06) | 0.001 |
| 1. Problems with getting along with other children | 53.93<br>(6.10) | 50.00<br>(5.92) | 60.95<br>(11.16) | 50.83<br>(2.12) | 0.004 |
| 2. Problems with other kids not wanting to be his or her friend | 45.37<br>(5.50) | 41.99<br>(4.31) | 42.41<br>(4.42) | 51.72<br>(2.05) | 0.333 |
| 3. Problems with getting teased by other children | 69.56<br>(3.49) | 65.70<br>(11.61) | 70.48<br>(13.21) | 72.5<br>(5.15) | 0.414 |
| 4. Problems with not being able to do things other children his or her age can do | 9.42<br>(4.48) | 11.49<br>(1.03) | 4.28<br>(0.40) | 12.5<br>(8.67) | 0.001 |
| 5. Problems with keeping up when playing with other children | 20.90<br>(9.55) | 23.09<br>(1.76) | 10.44<br>(1.96) | 29.16<br>(5.15) | 0.001 |
| **SCHOOL** | 40.39<br>(15.05) | 43.60<br>(16.34) | 38.57<br>(10.61) | 39.00<br>(18.78) | 0.063 |
| 1. Problems with paying attention in class | 27.58<br>(3.12) | 28.29<br>(3.16) | 30.29<br>(4.98) | 24.17<br>(5.10) | 0.001 |
| 2. Problems with forgetting things | 32.92<br>(5.12) | 33.58<br>(1.96) | 37.69<br>(5.74) | 27.5<br>(5.34) | 0.363 |
| 3. Problems with keeping up with schoolwork | 28.46<br>(4.75) | 33.89<br>(4.56) | 26.51<br>(3.53) | 25.00<br>(5.34) | 0.195 |
| 4. Problems with missing school because of not feeling well | 60.09<br>(7.47) | 64.65<br>(11.76) | 51.47<br>(6.30) | 64.17<br>(3.39) | 0.001 |
| 5. Problems with missing school to go to the doctor or hospital | 52.89<br>(5.45) | 57.59<br>(7.15) | 46.92<br>(6.33) | 54.17<br>(4.30) | 0.002 |

**Table 2.** FQOL Summary.

| **DOMAINS** | **Combined**<br>**Syndromic**<br>**ASDs N=391**<br>**(St. Dev.)** | **PMD**<br>**N=213**<br>**(St. Dev.)** | **RTT**<br>**N=148**<br>**(St.**<br>**Dev.)** | **SYNGAP1-**<br>**ID**<br>**N=30**<br>**(St. Dev.)** | **Kruskal-**<br>**Wallis H**<br>**Test**<br>**p-value** |
| --- | --- | --- | --- | --- | --- |
| **FAMILY INTERACTION** | 3.296<br>(0.940) | 3.33<br>(0.886) | 3.36<br>(0.970) | 2.69<br>(0.932) | 0.003 |
| 1. My family enjoys spending time together | 3.402<br>(1.303) | 3.385<br>(1.252) | 3.520<br>(1.353) | 2.933<br>(1.337) | 0.124 |
| 2. My family members talk openly with each other | 3.240<br>(1.264) | 3.371<br>(1.220) | 3.184<br>(1.282) | 2.600<br>(1.303) | 0.009 |
| 3. Our family solves problems together | 3.195<br>(1.233) | 3.288<br>(1.171) | 3.223<br>(1.282) | 2.400<br>(1.163) | 0.003 |
| 4. My family members support each other to accomplish goals | 3.189<br>(1.211) | 3.178<br>(1.200) | 3.311<br>(1.200) | 2.667<br>(1.241) | 0.038 |
| 5. My family members show that they love and care for each other | 3.742<br>(1.199) | 3.723<br>(1.163) | 3.905<br>(1.163) | 3.067<br>(1.413) | 0.153 |
| 6. My family is able to handle life's ups and downs | 3.003<br>(1.201) | 3.033<br>(1.203) | 3.061<br>(1.191) | 2.500<br>(1.167) | 0.075 |
| **PARENTING** | 3.04<br>(0.846) | 3.06<br>(0.847) | 3.11<br>(0.836) | 2.57<br>(0.769) | 0.004 |
| 7. My family members help the children be independent | 2.827<br>(1.227) | 2.920<br>(1.204) | 2.841<br>(1.217) | 2.100<br>(1.242) | 0.064 |
| 8. My family members help the children with schoolwork and activities | 2.807<br>(1.216) | 2.887<br>(1.222) | 2.767<br>(1.145) | 2.433<br>(1.455) | 0.167 |
| 9. My family members teach the children how to get along with each other | 3.189<br>(1.201) | 3.175<br>(1.208) | 3.315<br>(1.190) | 2.667<br>(1.093) | 0.087 |
| 10. Adults in our family teach the children to make good decisions | 3.348<br>(1.172) | 3.343<br>(1.156) | 3.428<br>(1.159) | 3.000<br>(1.313) | 0.554 |
| 11. Adults in my family know other people in my children's lives (friends, teachers, etc.) | 3.209<br>(1.292) | 3.192<br>(1.283) | 3.310<br>(1.288) | 2.833<br>(1.341) | 0.084 |
| 12. Adults in my family have time to take care of the individual needs of every child | 2.881<br>(1.291) | 2.850<br>(1.280) | 3.021<br>(1.310) | 2.433<br>(1.194) | 0.015 |
| **EMOTIONAL WELL-BEING** | 2.64<br>(0.904) | 2.67<br>(0.931) | 2.67<br>(0.895) | 2.266<br>(0.659) | 0.063 |
| 13. My family has the support we need to relieve stress | 2.584<br>(1.289) | 2.547<br>(1.274) | 2.748<br>(1.302) | 2.033<br>(1.189) | 0.120 |
| 14. My family members have friends or others who provide support | 2.700<br>(1.215) | 2.708<br>(1.250) | 2.689<br>(1.154) | 2.700<br>(1.291) | 0.352 |
| 15. My family members have some time to pursue our own interests | 2.522<br>(1.318) | 2.613<br>(1.346) | 2.524<br>(1.281) | 1.867<br>(1.137) | 0.075 |
| 16. My family has outside help available to us to take care of special needs of all family members | 2.767<br>(1.351) | 2.831<br>(1.314) | 2.735<br>(1.386) | 2.467<br>(1.432) | 0.062 |
| **PHYSICAL/MATERIAL WELL-BEING** | 3.67<br>(0.930) | 3.741<br>(0.903) | 3.59<br>(0.963) | 3.43<br>(0.9305) | 0.177 |
| 17. My family gets medical care when needed | 3.828<br>(1.211) | 3.882<br>(1.191) | 3.764<br>(1.258) | 3.767<br>(1.135) | 0.234 |
| 18. My family has dental care when needed | 3.723<br>(1.299) | 3.849<br>(1.226) | 3.547<br>(1.387) | 3.700<br>(1.291) | 0.164 |
| 19. My family members have transportation to get to the places they need to be | 3.549<br>(1.325) | 3.646<br>(1.289) | 3.439<br>(1.386) | 3.400<br>(1.248) | 0.334 |
| 20. My family has a way to take care of our expenses | 3.296<br>(1.303) | 3.363<br>(1.322) | 3.260<br>(1.254) | 3.000<br>(1.390) | 0.231 |
| 21. My family feels safe at home, work, school, and in our neighborhood | 3.931<br>(1.164) | 3.948<br>(1.174) | 3.973<br>(1.140) | 3.600<br>(1.192) | 0.370 |
| **DISABILITY RELATED SUPPORT** | 3.11<br>(0.991) | 3.1470<br>(0.316) | 3.13<br>(1.031) | 2.79<br>(1.018) | 0.202 |
| 22. My family member with a disability has support to accomplish goals at school or at workplace | 3.244<br>(1.259) | 3.394<br>(1.290) | 3.112<br>(1.175) | 2.793<br>(1.292) | 0.015 |
| 23. My family member with a disability has support to accomplish goals at home | 3.216<br>(1.270) | 3.268<br>(1.266) | 3.221<br>(1.250) | 2.833<br>(1.367) | 0.257 |
| 24. My family member with a disability has support to make friends | 2.784<br>(1.276) | 2.739<br>(1.251) | 2.885<br>(1.312) | 2.600<br>(1.276) | 0.502 |
| 25. My family has good relationships with the service providers who provide services and support to our family members with a disability | 3.324<br>(1.307) | 3.303<br>(1.328) | 3.419<br>(1.278) | 3.000<br>(1.287) | 0.207 |

## RESULTS

The PedsQL survey results revealed significant differences in health-related quality of life among the three syndromic autisms in each of the dimensions tested and in the overall score (Table 1 and Figure 1A). RTT girls had the lowest total HQoL score among our syndromic ASDs (M=38.03), followed by *SYNGAP1-*ID (M=43.51), and PMD (M=46.87). Across dimensions, the greatest impairment in syndromic ASDs was in physical functioning (M=33.55) (Figure 1B). When parsing the data based on genetic diagnosis, the greatest impairment for PMD was social functioning (M=38.45), while the greatest impairment for RTT was physical functioning (M=19.64) (Figure 1C and Table 1).

**FIGURE 1.**
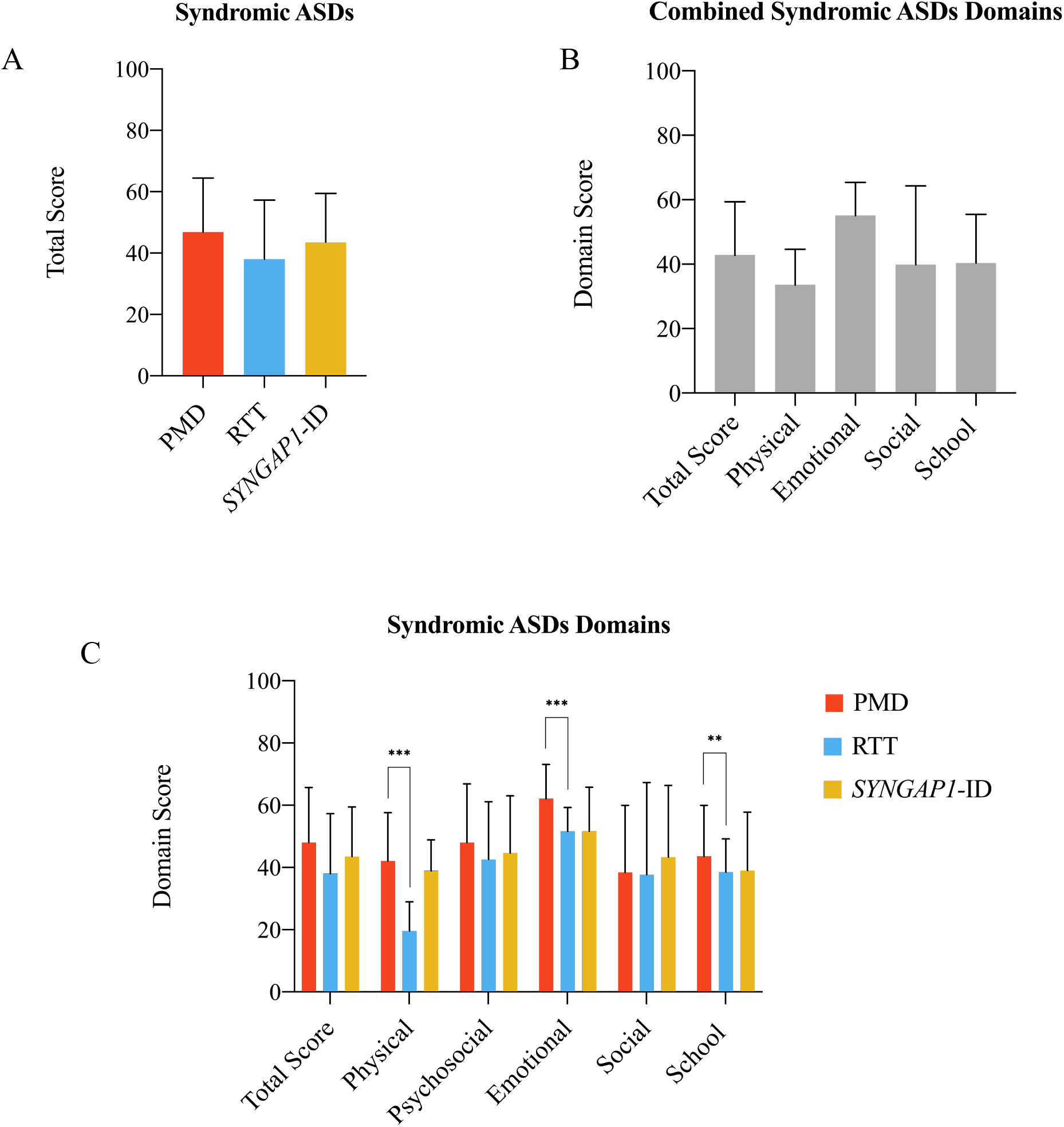
PedsQL 4.0 data from Phelan-McDermid syndrome, Rett syndrome, and *SYNGAP1*-related intellectual disability. A) Combined total scores for PMD, RTT and *SYNGAP*1-ID. B) Total score and individual domain scores for combined syndromic autism spectrum disorders. C) Total score and individual domain scores for PMD, RTT and *SYNGAP1*-ID. ** p<0.01, *** p<0.001 (student’s t-test).

For FQOL (Table 2 and Figure 2), the average scores of FQOL for combined syndromic autism for the family interaction (M=3.75), parenting (M=3.52) and disability-related support (M=3.64) were between satisfied and neither satisfied or dissatisfied. For emotional well-being, the average score for combined syndromic autism was between dissatisfied and neither satisfied or dissatisfied (M=2.79). Finally, for physical/material well-being, the average score for combined syndromic autism was in the satisfied range (M=4.05). Thus, the greatest toll on family quality of life for syndromic autism is on emotional well-being. There were significant differences among the syndromic autisms for parenting and family interaction which were driven by lower scores in the *SYNGAP1*-ID families. Direct comparison between RTT and PMD families revealed no significant differences for genetically-defined syndromic autisms demonstrating each has a similar impact on family quality of life.

**FIGURE 2.**
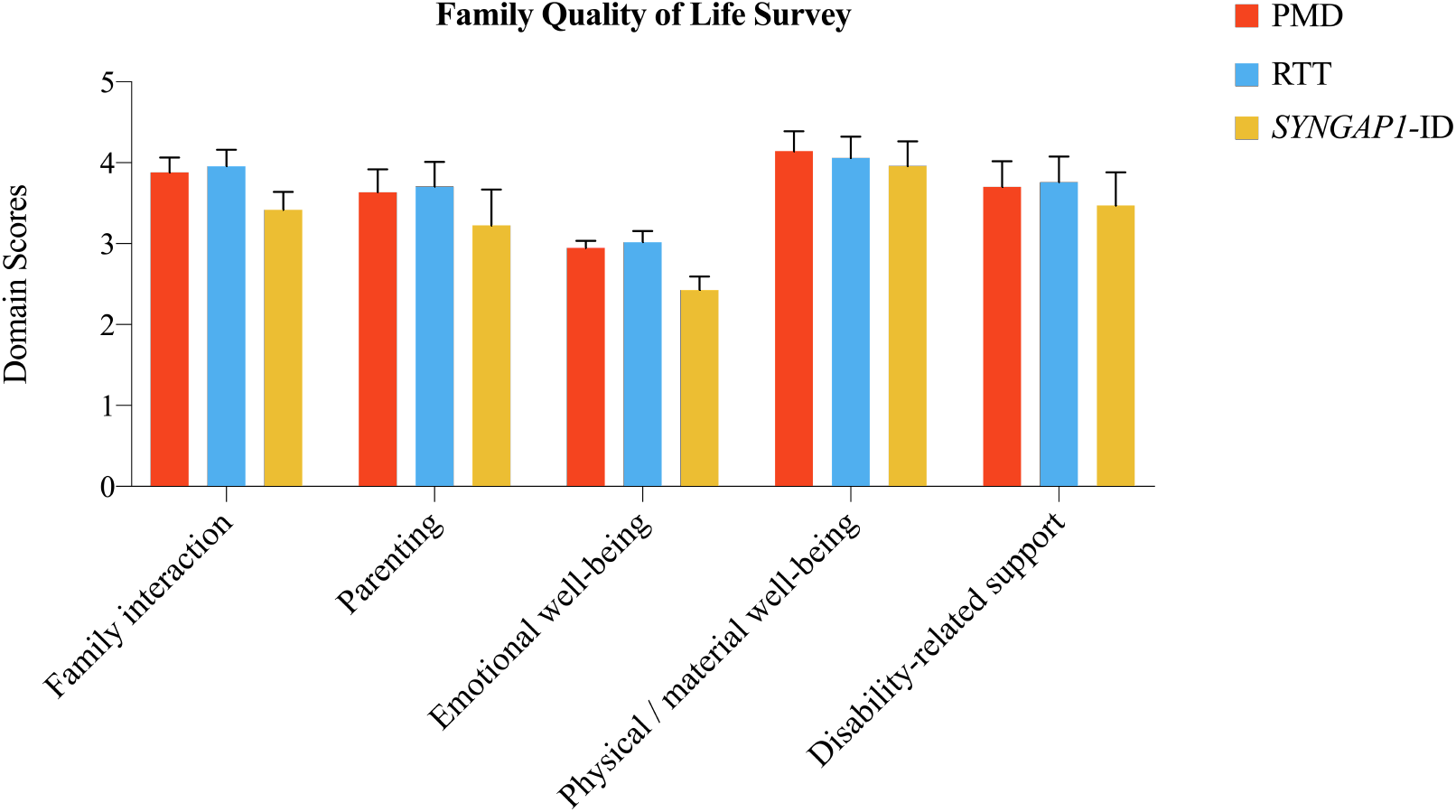
Beach Center Family Quality of Life Scale domain scores.

We next directly evaluated for differences between PMD and RTT patients in the PedsQL dimensions (Figure 1C). Overall, RTT patients scored worse than PMD across physical and emotional dimensions (p<0.001). However, no significant difference was identified in school or social domains. Thus, these data demonstrate significant differences among syndromic autisms in HRQoL of children.

We next assessed the relationship between each of the PedQL dimensions using correlation analysis and tested for inter-item reliabilities for each factor using Cronbach’s alpha. We found a good level of reliability (Tables 3-5) with physical and emotional functioning scores having the highest reliability. Furthermore, for PMD patients, the highest correlations were between physical and school domains as well as social and school domains. In contrast for RTT patients, the correlations across domains were generally lower compared to PMD patients. The highest correlations for RTT were between the physical and school domains as well as between emotional and school domains. The high correlation between physical and school domains likely reflects difficulty in school participation due to physical impairment in girls with RTT.

**Table 3.**
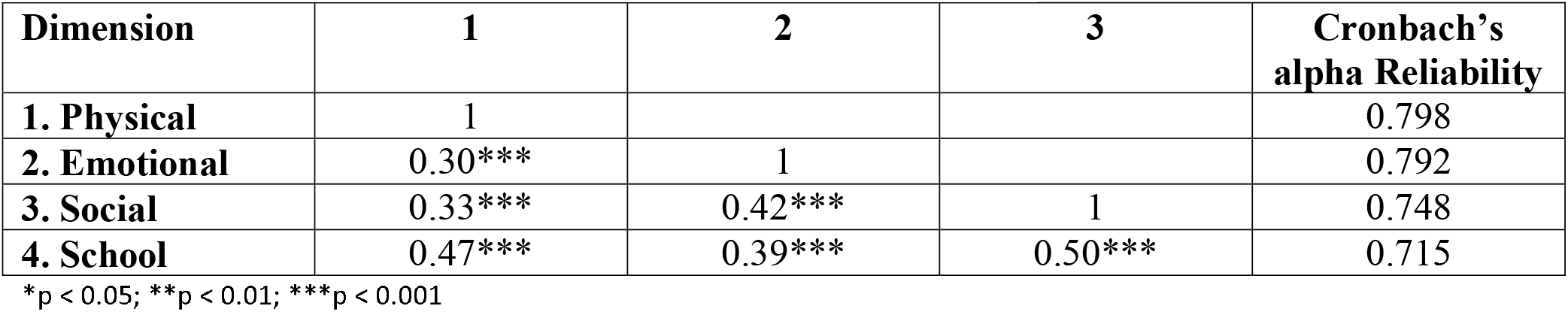
PedsQL correlations and inter-item reliabilities across all study measures and subscales PMD patients.

**Table 4.** PedsQL correlations and inter-item reliabilities across all study measures and subscales RTT patients.

| Dimension | 1 | 2 | 3 | Cronbach's alpha Reliability |
| --- | --- | --- | --- | --- |
| 1. Physical | 1 |  |  | 0.65 |
| 2. Emotional | 0.110*** | 1 |  | 0.67 |
| 3. Social | 0.273*** | 0.32*** | 1 | 0.68 |
| 4. School | 0.37*** | 0.25*** | 0.29*** | 0.67 |

\*p < 0.05; \*\*p < 0.01; \*\*\*p < 0.001

**Table 5.** PedsQL correlations and inter-item reliabilities across all study measures and subscales PMD and RTT patients.

| Dimension | 1 | 2 | 3 | Cronbach's alpha Reliability |
| --- | --- | --- | --- | --- |
| 1. Physical | 1 |  |  | 0.75 |
| 2. Emotional | 0.31*** | 1 |  | 0.75 |
| 3. Social | 0.29*** | 0.38*** | 1 | 0.73 |
| 4. School | 0.43*** | 0.37*** | 0.42*** | 0.68 |

\*p < 0.05; \*\*p < 0.01; \*\*\*p < 0.001

We next conducted principal components analyses for children’s PedsQL and caregivers’ FQOL (Table 6) separately by diagnosis and with all diagnoses combined. This analysis indicated that the number of factors for PedsQL and FQOL data matrices could be reduced into four to six factors and the percentage of explained variance was between 55% and 73%. This suggests that these individual dimension scores contain information, which might be truly heterogeneous in its nature for RTT and PMD patients and requires further investigation.

**Table 6.** Principal Component Analysis.

|  | Number of Factors with Eigenvalue >1 | Cumulative Variance Explained |
| --- | --- | --- |
| PedsQL PMD | 6 | 67% |
| PedsQL RTT | 6 | 73% |
| PedsQL PMD and RTT | 5 | 64% |
| FQOL PMD | 6 | 62% |
| FQOL RTT | 4 | 55% |
| FQOL PMD and RTT | 5 | 56% |

PedsQL 4.0 scores were compiled from published datasets in which similar age of inclusion criteria was implemented. Raw PedsQL 4.0 score data from healthy controls, type 1 (T1D) and type 2 diabetes (T2D)[18], intellectual disability (ID)[19], and idiopathic autism[12] were utilized as comparative assessment metric. For ease of interpretability, our PMD, RTT, and *SYNGAP1-*ID data were aggregated to create a new syndromic ASD cohort. A comparison of these total PedsQL cumulative scores across published datasets (Figure 3A) revealed highest scores in HRQoL among healthy individuals (M = 87.2), followed by type I diabetes (M = 76.61), type II diabetes (M = 74.36), autism (M = 65.2), ID (M = 56.62), and lowest in our syndromic ASDs (M = 42.80) (p = 0.0004). In comparing each dimension measured by the PedsQL among the chronic childhood disorders, syndromic autism consistently scored lower than other chronic childhood illnesses (Figure 3B).

**FIGURE 3.**
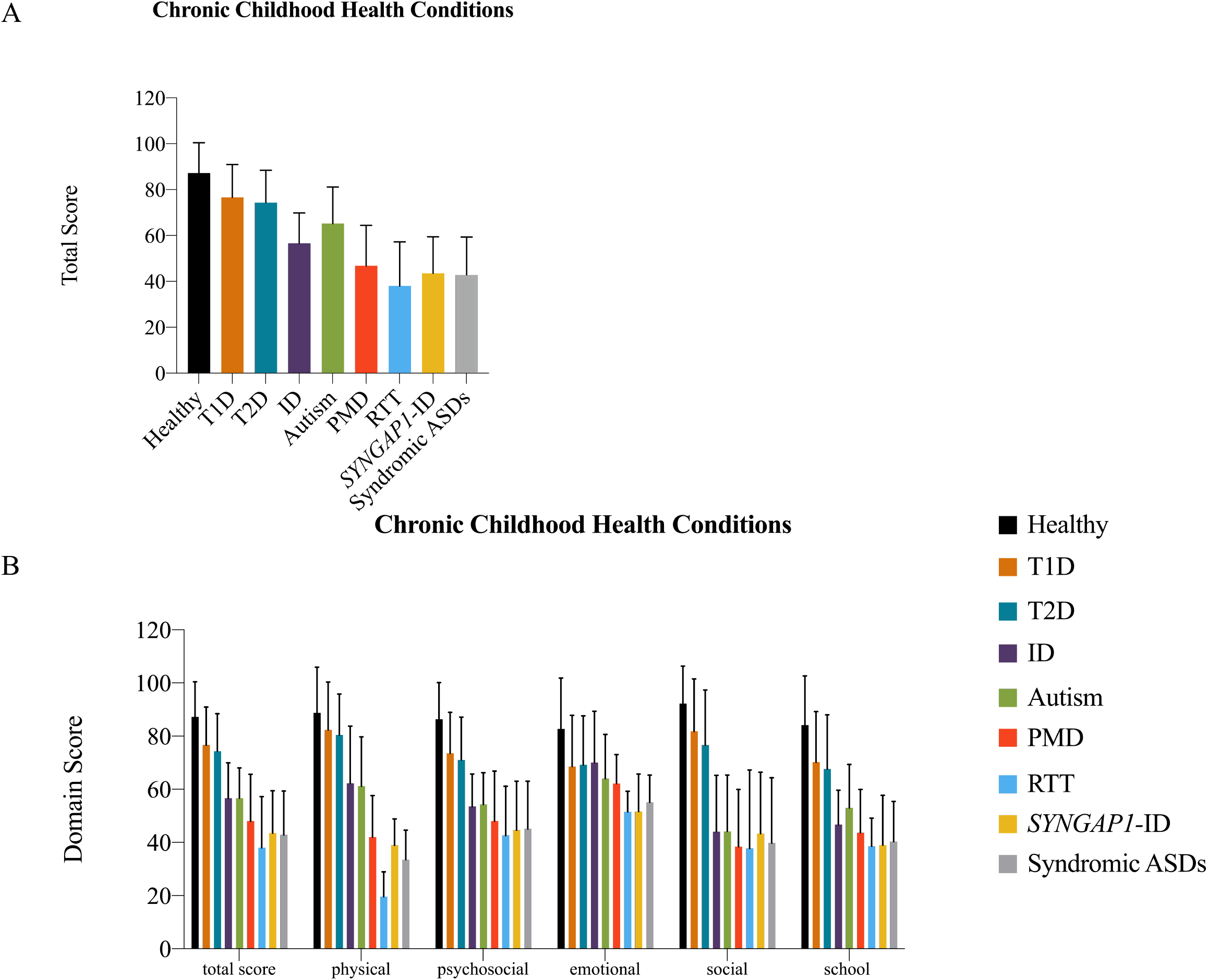
PedsQL of syndromic autism compared with other chronic childhood illnesses. A) Total quality of score and B) Domain scores for PedsQL across chronic childhood disorders.

## DISCUSSION

Through a validated and simple to administer survey, we discovered that children with syndromic autism spectrum disorders have significantly lower health-related quality of life compared with their neurotypical peers or children with other chronic health disorders. Among the syndromic autisms surveyed, we found that Rett syndrome had the lowest overall health-related quality of life based upon the total score from the PedsQL 4.0 survey driven primarily by lower physical functioning. Surprisingly, each syndromic autism child had a lower HRQoL scores than a idiopathic, non-syndromic autism child.

When comparing individual domains of the syndromic autism spectrum disorders with previously published data for idiopathic autism, we found similar scores in emotional, social and school performance as possibly expected due to both of these groups having clinical diagnosis of autism. Interestingly, the most significant difference between syndromic and idiopathic autism lies in the physical domain measured by the PedsQL. While this is driven by a significant degree by Rett syndrome, we do see greater reported impairment for Phelan-McDermid syndrome and *SYNGAP1*-ID in physical functioning compared with idiopathic autism as well.

We also found that ASDs not only affect the individual, but the entire family. Children with an ASD tend to report higher anxiety levels than neurotypical children and exhibit greater prevalence of co-occurring internalizing (depressed mood and anxiety) and externalizing (hyperactivity and aggression) behavior problems[20]. There is an elevated risk for parents (caregivers) of children with an ASD to experience mental health problems, such as stress, anxiety, and depression, compared to parents of children without an ASD or the general population. From clinical and transactional models, we know that there is a continuous reciprocal interaction process between children and caregivers. ASD symptoms, including behavioral dysregulation, contribute to increased stress in the parent (caregiver), which may impact the parents’ QOL and inadvertently alters parenting behaviors in ways that reinforce the child’s behavior problems and ASD symptoms, which can further influence the child’s QOL[20]. By viewing this from a transactional family dynamics perspective, we can see that ASDs not only affect the child but the whole family. Further studies addressing areas of improvement for families of children with syndromic ASDs regarding access to medical care, along with the support received by individuals with ASD may prove effective in addressing FQOL improvement.

Annual medical expenditures for children with ASD are four to six times greater than neurotypical children. It has been reported that the additional mean costs of caring for children diagnosed with ASD, including health care, education, ASD-related therapy, family-coordinated services and caregiver time total $17,081 per year[21]. Applying these estimates to the projected 1.7% of children aged 3 to 17 years with ASD in the U.S. results in a total societal cost of $268 billion as of 2015. The societal cost is forecasted to increase to $461 billion by the year 2025[22]. Similar economic modeling has not been performed for syndromic autisms. Given the greater concerns for health-related quality of life in syndromic autism compared with idiopathic autism, the costs per family associated with caring for children with syndromic autism spectrum disorders is presumptively even greater.

This work will aid in identifying therapeutic endpoints for evaluating treatment and intervention efficacy. This is true of both targeted therapy modalities such as physical, language and occupational therapies as well as therapies specifically targeting core autism symptoms such as Applied Behavior Analysis (ABA). From our data, physical impairments are a significant source of reduced quality of life for children with syndromic autism that is potentially overlooked in routine care. This research provides critical evidence, which might inform policymaking decisions regarding reconsideration of existing interventions aimed at patients with syndromic autism.

In addition to aiding clinicians in referring children with syndromic autism to established therapeutic interventions, this work provides a framework for future clinical trials. Many clinical trials for neurodevelopmental disorders have failed in recent years[23]. One hypothesis regarding these failures is that previous trials have been hampered with clinical endpoints (i.e., measurements of cognition) that are difficult to attain improvement in the limited timeframe of typical clinical trials. Health-related quality of life instruments provide a more holistic measurement of child health for clinical trials, that are arguably more robust than any instrument that measures a single neurodevelopmental variable.

This work has limitations that require future investigation. First, because of the anonymous nature of this study, the genetic diagnoses of participants could not be verified. Second, we focused on three genetic diagnoses that strongly predispose to syndromic autism. Whether this is representative of all syndromic autisms is unclear and warrants further investigation. Third, age was not collected in this online survey and as such could not be evaluated as a variable in the HRQoL scores. Future studies investigating correlation of HRQoL with additional co-variables such as age, congenital malformations, epilepsy and degree of intellectual disability will be informative.

## Data Availability

raw data is available upon request

## ACKNOWLEDGEMENTS

We are grateful to all of the families that participated in this study. We are also deeply indebted to the patient advocacy foundations that aided in subject recruitment including the Phelan-McDermid Syndrome Foundation, Bridge the GAP: *SYNGAP1* Education and Research Foundation, and RettSyndrome.org. We are grateful to Daniel Shallcross who helped set up the on-line survey. We are grateful to Claire de Oliveira for suggestions and comments.

## Abbreviations

HRQoL: health-related quality of life
FQOL: family quality of life
RTT: Rett syndrome
PMD: Phelan-McDermid syndrome
*SYNGAP1*-ID: *SYNGAP1*-related intellectual disability
ASD: autism spectrum disorder

## APPENDIX

### Phelan-McDermid syndrome

Phelan-McDermid syndrome (PMD) is due to either a chromosome 22q13 microdeletion or *SHANK3* loss-of-function mutations. PMD is characterized by developmental delay, impaired speech, dysmorphic features, intellectual disability, epilepsy and occasionally congenital cardiac or renal abnormalities. It is one of the most common monogenic forms of ASD[24] with approximately 84% of children with PMD fulfilling criteria for ASD[25]. Although the exact prevalence of PMD is unknown, at least 1,200 cases have been reported world-wide according to the Phelan-McDermid Syndrome Foundation, and PMD is considered to be a relatively common cause of intellectual disability, accounting for between 0.5% to 2.0% of cases.

Onset of symptoms occur during the second year of life with delays in acquisition of language and social skills. While developmental delays may not be apparent in the first 12 months of life, hypotonia is often present in infants, contributing to poor feeding, weak cry, and poor head control. Regression, the loss of acquired skills, is a recognizable feature of PMD[26] with regression of language being most common. Epilepsy is diagnosed in over 30% of children with PMD with a wide spectrum of severity[27]. No quantitative evaluation of the impact of this syndromic autism on health-related or family quality of life has ever been undertaken.

### Rett syndrome

Rett syndrome (RTT) is an X-linked dominantly inherited, progressive neurodevelopmental disorder, characterized by apparently normal psychomotor development during the first 6 to 18 months of life, followed by regression that affects speech, motor skills, and purposeful hand function[28]. It is the second leading cause of intellectual disability in girls, with an incidence ranging from 1/10,000 to 1/15,000 females born[29]. About 80% of females with classic RTT and 25 to 75% of those with variant forms, depending on the criteria used for diagnosis and size and age of sampled population, have mutations in *MeCP2* (methyl CpG binding protein 2), which encodes a chromatin binding protein[30]. The most recently revised clinical diagnostic criteria include partial or complete loss of acquired purposeful hand skills, regression of spoken language, onset of gait abnormalities which can include impaired (dyspraxia) or absence of ability (apraxia), and stereotypies including hand wringing/squeezing, clapping/tapping, mouthing, and washing/rubbing automatisms[31]. Additional characteristic features include postnatal deceleration of head growth, motor abnormalities, gait posture ataxia, breathing dysfunction, with seizures and breathing disturbances being the most detrimental clinical phenotype in RTT[32].

Recent studies have evaluated the QOL in subjects with RTT and their caregivers using questionnaires such as the Italian version of the Impact of Childhood Illness Scale[33]. These studies have demonstrated a significant negative impact on both the child’s development and the entire family.

### SYNGAP1 related intellectual disability

*SYNGAP1-*related intellectual disability (*SYNGAP1-*ID) is a neurological disorder characterized by moderate to severe intellectual disability evident in early childhood first described a decade ago. Early features consist of delayed speech and motor skills, with individuals typically having weak muscle tone (hypotonia), contributing greatly to difficulties with motor development[34]. Autism spectrum disorder is diagnosed in as many as 73% of individuals with pathogenic *SYNGAP1* mutations[35]. Moreover, in unbiased exome sequencing of over eleven thousand individuals with autism, *SYNGAP1* mutations were among the top three monogenic causes of autism[6]. Early features of *SYNGAP1*-ID are usually present within the first two years of life, and typically include severely delayed development of language and motor skills. Children are often either non-verbal or minimally verbal. On average, they walk independently for the first time on average at 22 months of age, which is approximately double that of neurotypical children. Behavioral abnormalities may include hand flapping, obsessions with certain objects and poor social development. Other behavioral abnormalities include hyperactivity, impulsivity, physical aggression, mood swings, sullenness and rigidity. Greater than 90% of children with *SYNGAP1*-related intellectual disability have epilepsy which can be intractable and devastating[35, 36]. To date, no studies investigating the impact on health-related or family quality of life have been conducted for *SYNGAP1*-related intellectual disability.

## Notes

**Funding source:** This study was supported by the Robbins Foundation (to Drs. Bolbocean and Holder). Dr. Holder is also generously supported by the Joan and Stanford Alexander Endowed Chair and Mr. Charif Souki.

**Conflict of interest:** All authors have no potential conflicts of interest to disclose.

### Competing Interest Statement

The authors have declared no competing interest.

### Funding Statement

This study was supported by the Robbins Foundation (to Drs. Bolbocean and Holder). Dr. Holder is also generously supported by the Joan and Stanford Alexander Endowed Chair and Mr. Charif Souki.

### Author Declarations

Baylor College of Medicine Institutional Review Board approved this work.

